# The Impact of Treatment Frequency on Therapeutic Alliance and Clinical Outcomes in Acupuncture for Gulf War Illness

**DOI:** 10.1101/2025.04.22.25326212

**Authors:** L.M. Johnson, A Incollingo-Rodriguez, B. Nephew, L. Conboy

**Affiliations:** University or Utah; Worcester Polytechnic Institute; Harvard Medical School

**Keywords:** Therapeutic Alliance, Gulf War Illness, Acupuncture, Treatment Frequency, Working Alliance Inventory (WAI-SR)

## Abstract

**Purpose:** This study evaluates the development of therapeutic alliance (TA) and its clinical influence on acupuncture treatments for Gulf War Illness (GWI).

**Methods:** Data were collected from a 3.5-year US Army-funded randomized clinical trial (RCT) involving thirty-two experienced acupuncturists who provided individualized treatments over approximately six months. To measure improvements, paired-sample t-tests compared Working Alliance Inventory (WAI-SR) scores across task, bond, and goal factors from baseline to endpoint (6 months). Hierarchical linear regression analyses assessed the mediating effects of therapeutic alliance variables on pain and functional outcomes at the six-month mark.

**Results:** Both groups had similar WAI-SR scores at the endpoint (6 months); however, initial scores in all three factors were lower in the low-dose (weekly acupuncture) group compared to the higher-dose (biweekly acupuncture) group. The weekly treatment group exhibited significant improvements in WAI-SR scores across all factors from baseline (2 months) to endpoint (6 months) (p < 0.01). Linear regression analyses indicated that biweekly treatments were associated with significantly lower pain levels at six months.

**Conclusion:** These findings suggest that the frequency of acupuncture treatments influences the development of the therapeutic alliance and clinical outcomes.

## 1. Introduction

The therapeutic alliance (TA) is a fundamental aspect of the patient-practitioner relationship, playing a critical role in achieving positive clinical outcomes. However, developing and maintaining these relationships can be challenging, highlighting the need for healthcare practitioners to have evidence-based strategies to foster positive interactions. Research over several decades has identified three essential components of TA as proposed by Edward S. Brodin (1979): agreement on goals (goals), agreement on interventions (tasks), and the quality of the bond between patient and practitioner (bond) [1–3]. Building on Brodin’s theory, Horvath and Goldberg (1989) developed the Working Alliance Inventory (WAI). This 36-item assessment tool evaluates TA on a 7-point scale, ranging from 1 (never) to 7 (always), based on self-reported patient and practitioner clinical outcomes [2]. The WAI can be adapted for various complementary medicine therapies, including acupuncture, massage, herbal medicine, and dietary therapy.

A qualitative study extracted data from a randomized controlled trial (RCT) to explore patient experiences with acupuncture for lower back pain (LBP) and the formation of TA. This study underscored the significance of the TA relationship in achieving self-reported outcomes, such as pain relief, improvements in physical activity, relaxation, psychological benefits, and reduced reliance on medication [4]. The formation of TA was identified as a driving force, enabling patients to express their concerns regarding barriers to treatment, including discomfort with needles, temporary symptom exacerbation, pressure to continue treatment, and financial constraints [4].

Another prospective cohort study gathered patient-reported and practitioner-reported data at various intervals, including baseline (first session), two weeks post-baseline, and three months post-baseline. The study involved 166 practitioners (including 55 acupuncturists) and 960 adult patients, focusing on mediators such as patient self-efficacy for management and perceptions of lower back pain and psychosocial distress [5]. The findings indicated robust TA components (goals, tasks, and bonds) contributed to increased patient satisfaction, reduced skepticism about treatment credibility, and heightened positive outcome expectancies among practitioners [5].

Clinical research supports the importance of TA in the practitioner-patient relationship as a driving force behind various outcomes, even in complex situations involving multiple and intricate symptoms or conditions [2, 6–22]. Consequently, healthcare practitioners must establish a foundation of trust and collaboration, balancing vulnerability and authority to foster realistic expectations. This approach enables patients to achieve their overall health goals and engage effectively with treatment tasks, leading to positive results and stronger bonds.

Given the complexity of TA, this study focuses on gathering, reviewing, and synthesizing data from randomized controlled trials examining individualized acupuncture sessions for veterans diagnosed with Gulf War Illness (GWI), a multifaceted diagnosis characterized by symptoms such as fatigue, sleep disturbances, mood disorders, cognitive dysfunction, and musculoskeletal pain [23, 24]. The aim is to investigate (1) the development of TA, (2) the timeframe for its formation, and (3) its effect on overall patient outcomes, particularly regarding symptom management (e.g., pain levels). Based on existing literature, the hypothesis posits that TA will increase over time due to concordance (agreement on treatment), resulting in overall patient pain reduction. Self-reported mediators, which are measurable factors influenced by the practitioner’s expertise, will also be considered. The data from this study aims to inform practitioners about the importance of cultivating patient-practitioner relationships and their influence on patients’ pre-, during, and post-treatment outcomes, encouraging practitioners to actively engage in achieving TA with all patients.

## 2. Methods

This study conducted a secondary data analysis from a 3.5-year, US Army-funded randomized clinical trial (RCT) approved by the New England Institutional Review Board (IRB # 09-204) and the U.S. Army Human Research Protection Office (Award No. W81XWH-09-2-0064). The analysis aimed to investigate the impact of the therapeutic alliance (TA) features, the timeline of TA development, and the components of concordance and mediation on the therapeutic outcomes of veterans who received acupuncture treatment for Gulf War Illness (GWI).

Specifically, the study examined the relationship between treatment conditions and TA levels of pain scores at six months.

### 2.1 Original GWI Army-Funded Acupuncture RCT Overview

The original US Army-funded RCT involved thirty-two experienced acupuncturists providing individualized treatments to 104 veterans diagnosed with GWI [25, 26]. All licensed acupuncturists had at least five years of clinical experience and received additional in-house training from the VA regarding GWI. Participants provided informed consent and were randomized into two groups: (a) biweekly acupuncture for six months (Group 1, n = 52) or (b) a 2-month waitlist, followed by weekly acupuncture for four months (Group 2, n = 52) [25, 26]. Each acupuncture intervention lasted approximately one hour and included an initial assessment to develop an individualized treatment plan. Acupuncture points were selected based on Traditional Chinese Medicine (TCM) principles and differential diagnosis, with needles retained for 30–45 minutes [25, 26]. Additional therapies, including electroacupuncture, heat therapies (e.g., heat lamps), Chinese massage (tui na), cupping, and applying press balls, tacks, or magnets to acupoints post-treatment, were permitted. However, herbal medicine and supplements were prohibited [25, 26]. Treatments were conducted in a private setting outside the Veteran Health Administration (VA) Hospital, and participants continued to receive standard care as necessary during the study.

### 2.2 Therapeutic Alliance Assessment Overview

The TA assessment in this study was based on the original Working Alliance Inventory (WAI) scale created by Horvath and Goldberg, modified with questions specific to acupuncture practitioners [19, 20]. The adjusted WAI was administered to patients at baseline and at 2, 4, and 6 months, regardless of their randomized control group assignment [17]. To further evaluate whether congruence in patient-practitioner relationships was indicative of superior treatment outcomes, Working Alliance Inventory-Short Revised (WAI-SR) scores were utilized and compared to clinical outcomes assessed through the Affective Social Support (Interpersonal Support Evaluation List) (ISEL sum) [28], Total Beck Depression score (CDEP Depression sum) [29]—total Beck Anxiety score (CDEP Psychological Anxiety sum) [30]. WAI-SR scores were analyzed for task and goal agreement and the formation of affective bonds during treatment.

### 2.3 Statistical Analysis

Average WAI scores were calculated for each participant and practitioner, and the patient-practitioner graphed results for each factor over time. WAI-SR scores were compared using paired sample Student’s t-tests throughout the study. The change in concordance for each patient-practitioner relationship was computed from baseline to the six-month endpoint. Linear regression models measured the influence of changes in concordance on pain and physical function outcomes. Hierarchical linear regression analyses tested the therapeutic alliance variables (Task, Bond, and Goal) as mediators in the relationship between treatment group assignment and pain and functional outcomes at six months, controlling for baseline pain and function levels. Statistical significance was set at p ≤ 0.05.

## 3. Results

The study demographics included participant population characteristics such as age, gender, and self-reported race. The mean age of participants was 48.2 years, with 14% identifying as female, 86% as male, and 81% self-reporting as white (see Table 1). No significant differences were observed between the two treatment groups.

**Table 1.** Baseline Characteristics of Study Population.

| Characteristics | Treatment |
| --- | --- |
| Age-year ± Standard deviation ($N = 103$) | 48.2 ± 7.5 |
| Female sex, $N$ (%) ($N= 104$) | 14 (13.5) |
| **Self-reported race, $N$ (% of total) ($N = 104$)** |  |
| White | 84 (80.8) |
| Black or African American | 10 (9.6) |
| Asian | 1 (1.0) |
| American Indian/Alaskan Native | 1 (1.0) |
| More than one race | 2 (1.9) |
| Unknown | 1 (1.0) |
| Other | 5 (4.8) |
| **Self-reported Hispanic, *N* (% of total) (*N* = 104)** |  |
| Yes | 6 (5.8) |
| **No** | 94 (90.4) |
| No Answer | 4 (3.8) |

As the study progressed, patient-practitioner pairs reported increasingly favorable scores across all factors of the Working Alliance Inventory-Short Revised (WAI-SR)—Task, Bond, and Goal (see Figure 1). The patient-practitioner relationships showed higher levels of concordance over time (see Figure 2). While both the weekly and biweekly groups had similar WAI-SR scores at the six-month endpoint, initial WAI-SR scores for all factors (Task, Bond, and Goal) were lower in the weekly group compared to the biweekly group. Notably, scores for all WAI-SR factors in the weekly treatment group increased more sharply than those in the biweekly group (see Figure 1). The weekly treatment group had two months on the waitlist before starting treatment, highlighting the importance of allowing sufficient clinical time for the patient-practitioner relationship to strengthen. Increased interaction and treatment frequency contributed to an improved therapeutic alliance.

**Figure 1.**
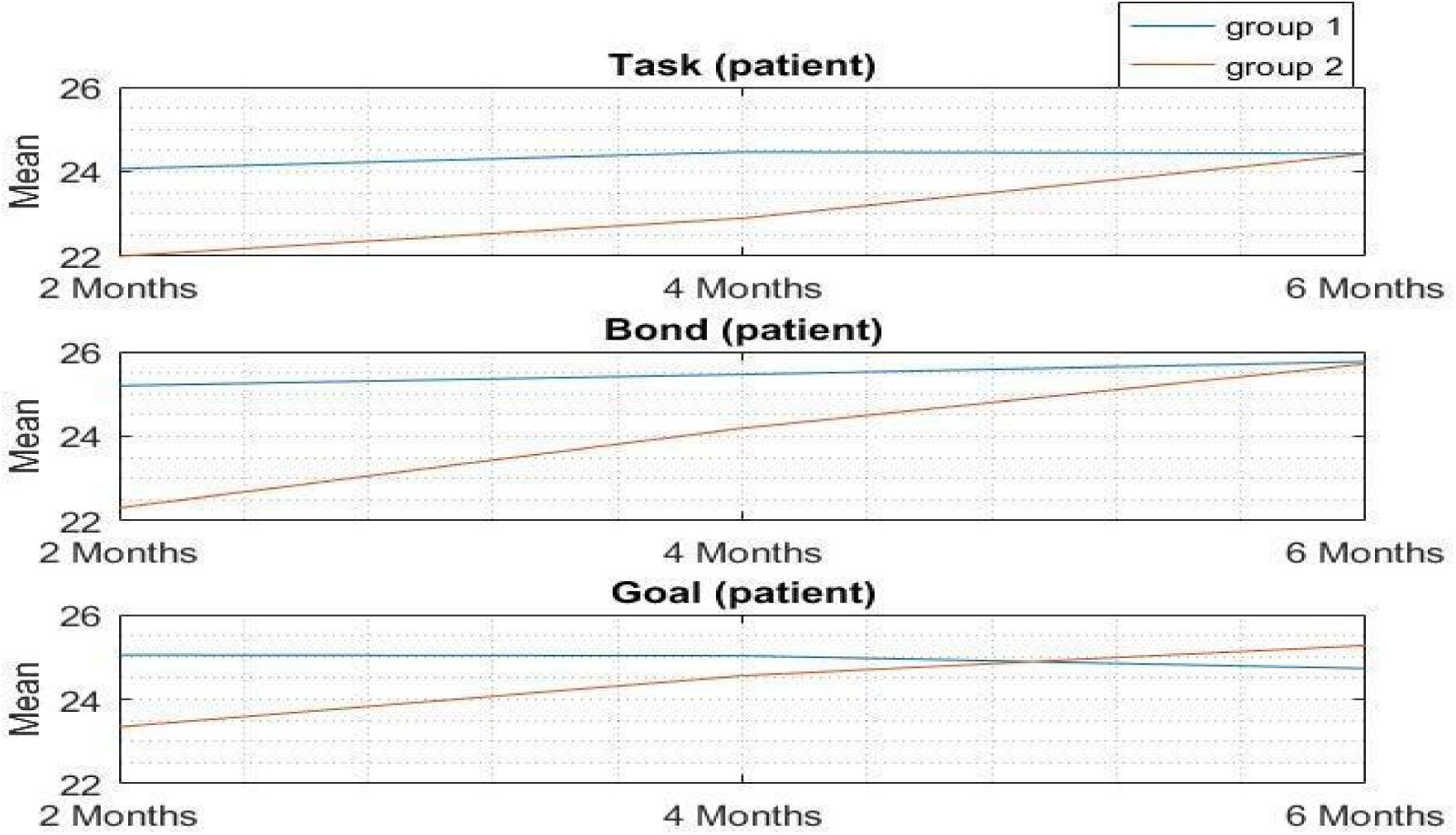
Patients’ WAI-SR scores versus times for groups 1 (biweekly) & 2 (weekly waitlisted).

**Figure 2.**
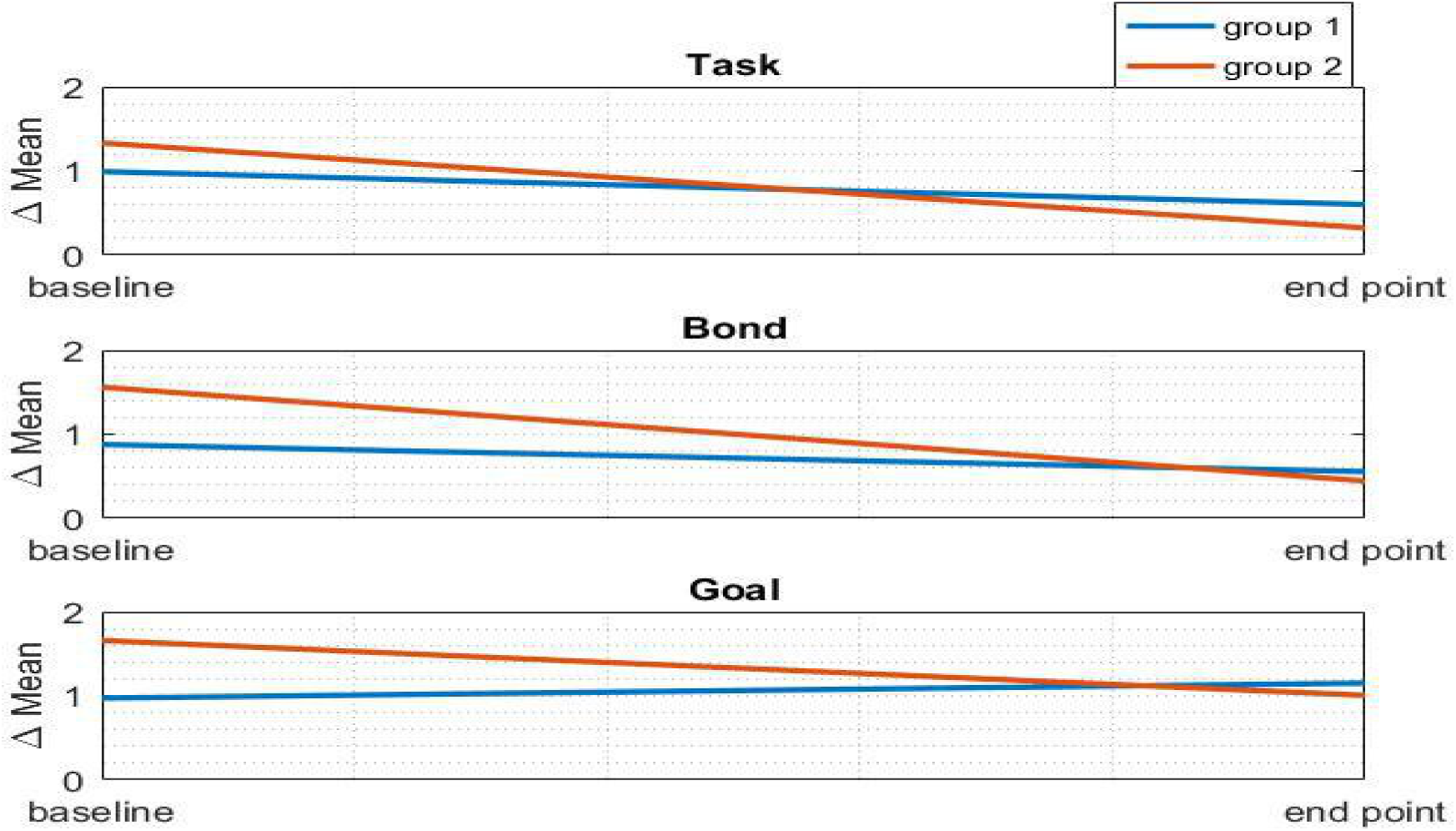
Δ Patient Practitioner WAI-SR scores versus times for groups 1 (biweekly) & 2 (weekly waitlisted).

**Figure 3.**
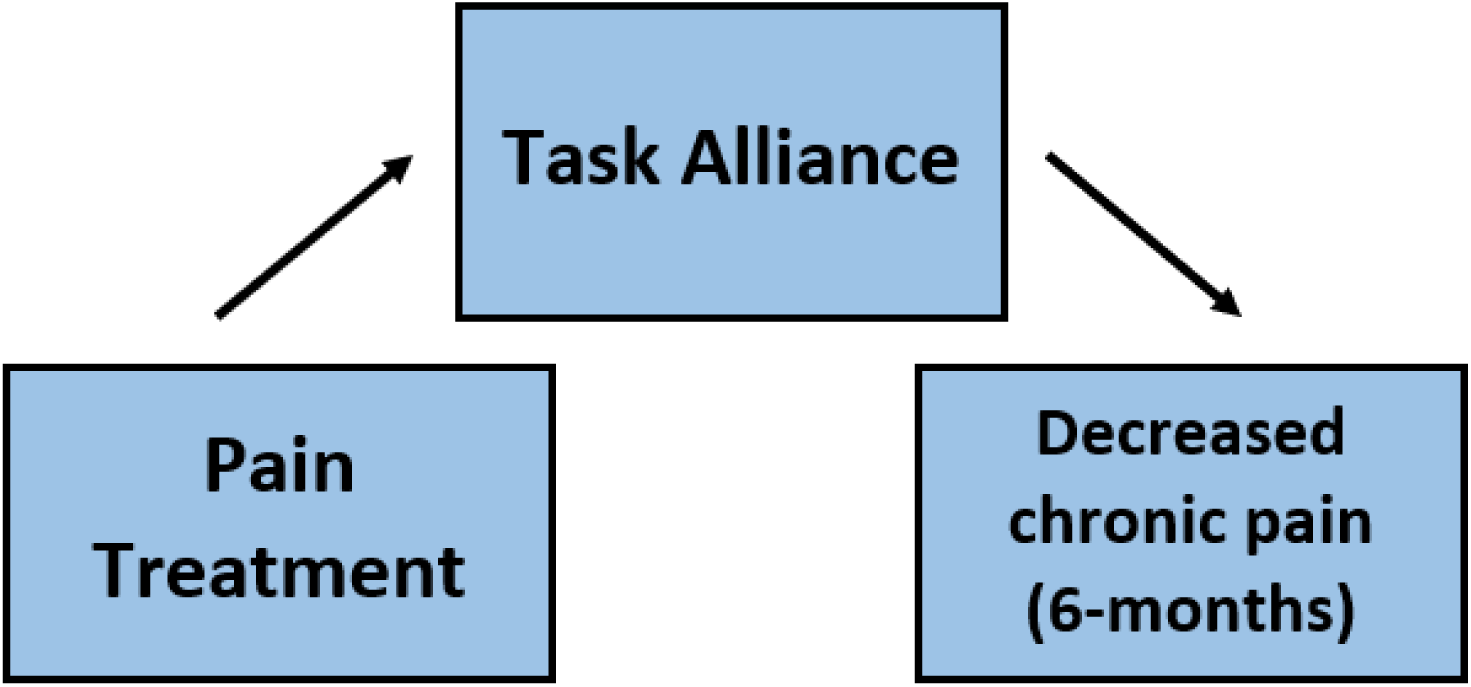
Summary of mediation for the treatment of pain.

Paired-sample t-tests indicated that WAI-SR scores for the weekly group showed significant improvement from baseline (2 months) to endpoint (6 months) across all factors (Task, Bond, and Goal) (see Tables 2-4).

**Table 2.** WAI-SR Scores in Task Factor.

| Pair | Time Point | Treatment | Mean | SD | N | p-Value | Diff. Means |
| --- | --- | --- | --- | --- | --- | --- | --- |
| Pair 1 | 2 months | Biweekly | 24.07 | 3.08 | 45 | 0.5858 | 0.4 |
|  | 4 months | Biweekly | 24.46 | 3.54 | 39 |  |  |
|  | 2 months | Weekly | 22 | 6.20 | 37 | 0.5186 | 0.9 |
|  | 4 months | Weekly | 22.9 | 5.75 | 38 |  |  |
| Pair 2 | 2 months | Biweekly | 24.07 | 3.08 | 45 | 0.5945 | 0.36 |
|  | 6 months | Biweekly | 24.43 | 3.24 | 42 |  |  |
|  | 2 months | Weekly | 22 | 6.20 | 37 | 0.0504 | 2.42 |
|  | 6 months | Weekly | 24.42 | 4.18 | 38 |  |  |
| **Pair 3** | 4 months | Biweekly | 24.46 | 3.54 | 39 | 0.9652 | 0.03 |
|  | 6 months | Biweekly | 24.43 | 3.24 | 42 |  |  |
|  | 4 months | Weekly | 22.9 | 5.75 | 38 | 0.1894 | 1.52 |
|  | 6 months | Weekly | 24.42 | 4.18 | 38 |  |  |

**Table 3.**
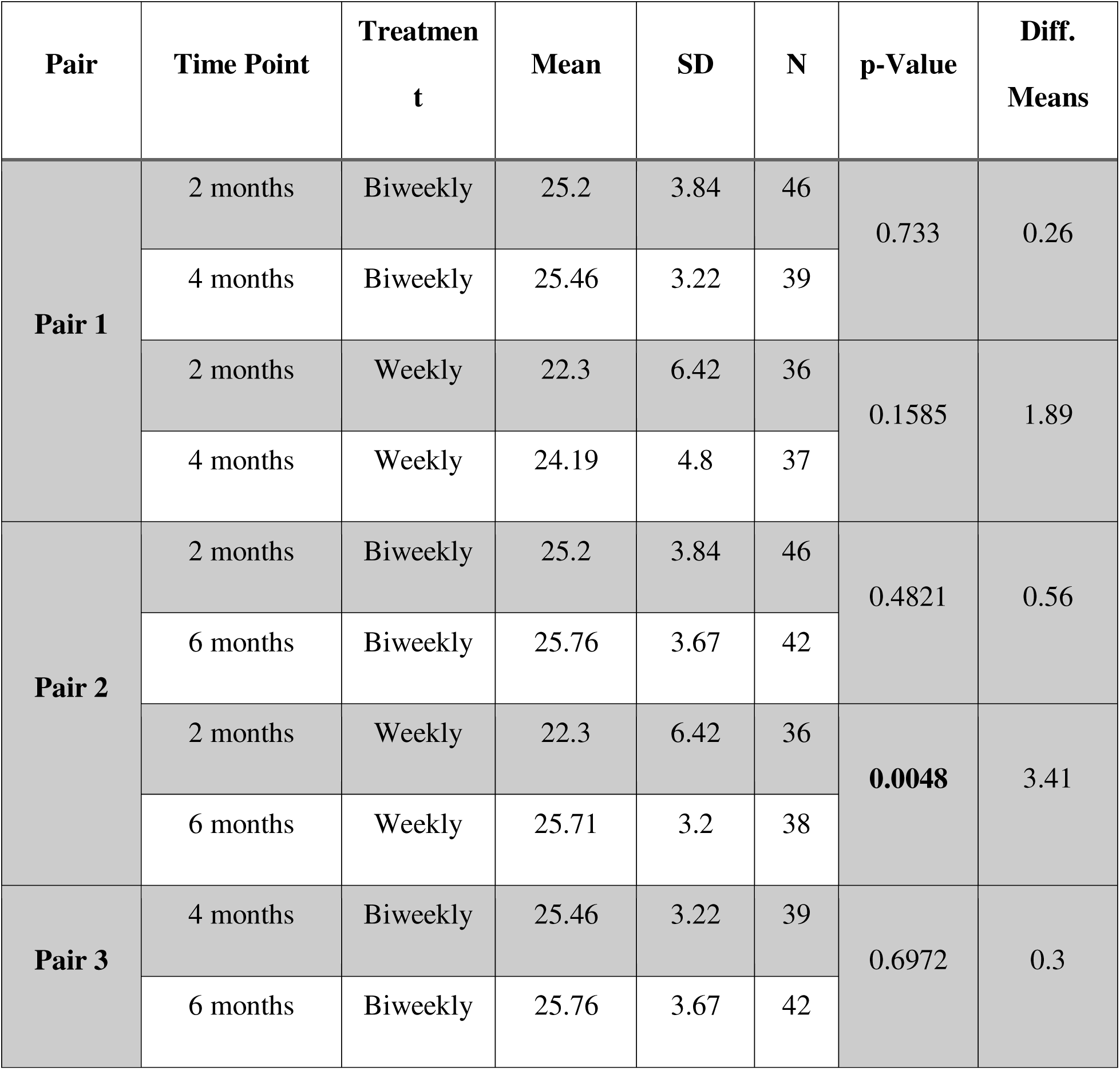
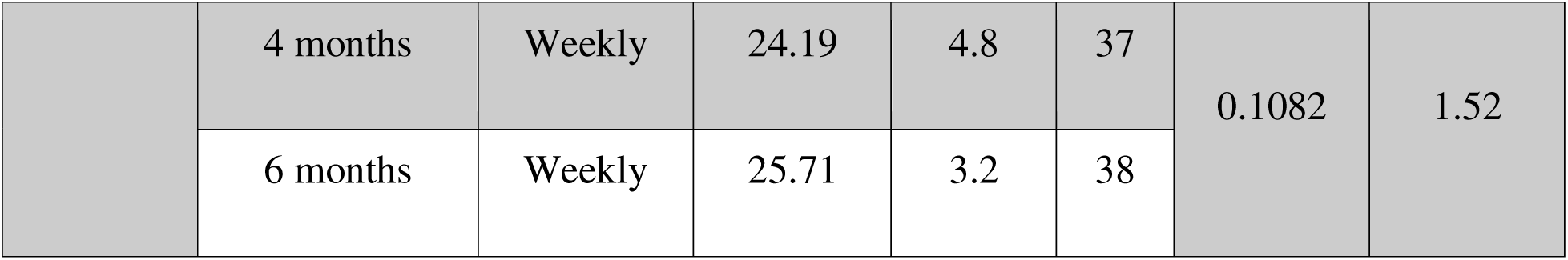
WAI-SR Scores in Bond Factor.

**Table 4.** WAI-SR scores in Goal factor.

| Pair | Time Point | Treatmen<br>t | Mean | SD | N | Diff.<br>Means | p-value |
| --- | --- | --- | --- | --- | --- | --- | --- |
| **Pair 1** | 2 months | Biweekly | 25.04 | 3 | 45 | 0.01 | 0.98 |
|  | 4 months | Biweekly | 25.03 | 3.57 | 38 |  |  |
|  | 2 months | Weekly | 23.34 | 3.69 | 35 | 1.21 | 0.1657 |
|  | 4 months | Weekly | 24.55 | 3.69 | 38 |  |  |
| **Pair 2** | 2 months | Biweekly | 25.04 | 3 | 45 | 0.31 | 0.6706 |
|  | 6 months | Biweekly | 24.73 | 3.92 | 42 |  |  |
|  | 2 months | Weekly | 23.34 | 3.69 | 35 | 1.93 | **0.0228** |
|  | 6 months | Weekly | 25.27 | 3.34 | 37 |  |  |
| **Pair 3** | 4 months | Biweekly | 25.03 | 3.57 | 38 | 0.3 | 0.7221 |
|  | 6 months | Biweekly | 24.73 | 3.92 | 42 |  |  |
|  | 4 months | Weekly | 24.55 | 3.69 | 38 | 0.72 | 0.3803 |
|  | 6 months | Weekly | 25.27 | 3.34 | 37 |  |  |

**Table 5.** Social Support, Depression, and Anxiety Correlations.

| **Treatment Biweekly (Group 1)** |  |  |  |  |  |  |
| --- | --- | --- | --- | --- | --- | --- |
| **Clinical Outcome** | **ΔTask** | **p-value** | **ΔBond** | **p-value** | **ΔGoal** | **p-value** |
| **Affective Social Support (N=36)** | -0.099 | 0.282 | 0.105 | 0.272 | 0.06 | 0.364 |
| **Δ Affective Social Support**<br>**(N=36)** | -0.051 | 0.385 | -0.012 | 0.472 | 0.149 | 0.193 |
| **Total Beck Depression score**<br>**(N=37)** | -0.16 | 0.173 | -0.21 | 0.106 | -0.071 | 0.338 |
| **Δ Total Beck Depression score**<br>**(N=37)** | 0.029 | 0.432 | 0.035 | 0.418 | 0.085 | 0.309 |
| **Total Beck Anxiety score (N=37)** | -0.08 | 0.318 | -0.205 | 0.112 | -0.107 | 0.265 |
| Δ Total Beck Anxiety score (N=37) | 0.147 | 0.193 | -0.02 | 0.453 | 0.117 | 0.246 |
| **Treatment Weekly Waitlisted (Group 2)** |  |  |  |  |  |  |
| **Clinical Outcome** | **ΔTask** | **p-value** | **ΔBond** | **p-value** | **ΔGoal** | **p-value** |
| Affective Social Support (N=36) | -0.21 | 0.125 | -0.081 | 0.33 | -0.196 | 0.141 |
| Δ Affective Social Support (N=36) | 0.092 | 0.308 | 0.114 | 0.267 | -0.037 | 0.421 |
| Total Beck Depression score<br>(N=37) | -0.044 | 0.406 | -0.062 | 0.367 | 0.107 | 0.281 |
| Δ Total Beck Depression score<br>(N=37) | 0.114 | 0.267 | -0.142 | 0.22 | 0.009 | 0.481 |
| Total Beck Anxiety score (N=37) | -0.214 | 0.12 | **-0.418** | **0.009** | -0.103 | 0.287 |
| Δ Total Beck Anxiety score (N=37) | 0.117 | 0.262 | -0.042 | 0.41 | 0.128 | 0.243 |

\*Affective Social Support (*ISEL sum*); Total Beck Depression score (*CDEP Depression sum*);
Total Beck Anxiety score (*CDEP Psychological Anxiety sum*).

**Table 6.** Overall Clinical Outcome Correlations.

|  |  |  |  |  |  |  |
| --- | --- | --- | --- | --- | --- | --- |
| **Treatment Biweekly (Group 1)** |  |  |  |  |  |  |
| **Clinical** | **Task** | **p-value** | **Bond** | **p-value** | **Goal** | **p-value** |

| **Outcome** |  |  |  |  |  |  |
| --- | --- | --- | --- | --- | --- | --- |
| Δ SF-36 (N=37) | -0.166 | 0.162 | -0.269 | 0.054 | **-0.281** | **0.046** |
| Δ McGill (N=37) | 0.000 | 0.499 | -0.221 | 0.095 | 0.044 | 0.399 |
| **Treatment Weekly Waitlisted (Group 2)** |  |  |  |  |  |  |
| **Clinical Outcome** | **Task** | **p-value** | **Bond** | **p-value** | **Goal** | **p-value** |
| Δ SF-36 (N=32) | **-0.309** | **0.042** | -0.158 | 0.195 | **-0.345** | **0.027** |
| Δ McGill (N=32) | 0.126 | 0.250 | 0.144 | 0.220 | 0.062 | 0.371 |

Regression analyses revealed that concordance across subscales was associated with improved clinical outcomes. Notably, the Goal factor (shared treatment goals) was significantly related to improvements in physical functioning, as measured by the SF-36 physical function subscale, in both groups (p < 0.05). Higher concordance was associated with more significant improvements in SF-36 physical function.

Within the weekly treatment group, the Total Beck Anxiety Score significantly correlated with the Bond factor (p = 0.009) (see Table 5).

Although treatment conditions did not significantly predict practitioner therapeutic alliance variables, they did predict patient-reported therapeutic alliance variables. Further analyses tested these variables to mediate the relationship between treatment conditions and pain/function outcomes.

Treatment condition emerged as a significant predictor of pain levels on the McGill Pain Scale at six months (b =-3.77, SE = 1.63, p =.023), indicating that the biweekly treatment group (Group 1) reported significantly lower pain levels at six months. Results suggested that the Task factor substantially mediates the relationship between treatment and pain levels at six months.

Specifically, the treatment group significantly predicted task alliance levels, which, in turn, significantly predicted six-month pain levels. When accounting for task alliance, the direct path from treatment to six-month pain became non-significant, indicating complete mediation.

However, the Bond and Goal working alliance variables at two months did not significantly mediate the relationship between treatment condition and six-month pain levels. See **Table 7** for regression coefficients.

**Table 7.** Hierarchical Linear Regression Analyses Testing Mediators of Treatment to Pain.

| Task (*n* = 64) |  |  |  |  |
| --- | --- | --- | --- | --- |
| Path | *B* | *SE B* | $\beta$ | *p*-value |
| Treatment to task | 2.28 | 1.14 | .23 | **.049** |
| Task to 6-month pain (accounting for treatment) | -0.40 | 0.18 | -.22 | **.032** |
| Treatment to 6-month pain (controlling for the task) | -2.13 | 1.89 | -.11 | .263 |
| Bond (*n* = 64) |  |  |  |  |
| Path | *B* | *SE B* | $\beta$ | *p*-value |
| Treatment to bond | 3.04 | 1.24 | .28 | **.017** |
| Bond to 6-month pain (accounting for treatment) | -0.18 | 0.17 | -.11 | .310 |
| Treatment to 6-month pain (controlling for the bond) | -3.27 | 1.95 | -.18 | .099 |
| Goal (*n* = 63) |  |  |  |  |
| Path | *B* | *SE B* | $\beta$ | *p*-value |
| Treatment to goal | 1.66 | 0.80 | .24 | **.042** |
| Goal to 6-month pain (accounting for treatment) | -0.32 | 0.28 | -.12 | .258 |
| Treatment to 6-month pain (controlling for the goal) | -2.36 | 1.95 | -.13 | .232 |

Note. All models control for baseline pain.

## 4. Discussion

This study investigated the development, formation, and effects of therapeutic alliance (TA) on the pain outcomes of veterans diagnosed with Gulf War Illness (GWI) who completed individualized acupuncture sessions. Data indicated that all WAI-SR factors (Task, Bond, and Goal) exhibited positive scores that increased throughout the study, with both treatment groups achieving similar scores at the six-month endpoint. This finding is significant as it demonstrates the development of TA in both groups: (a) biweekly (twice per week) acupuncture for six months and (b) a two-month waitlist followed by weekly acupuncture for four months. The waitlist group exhibited lower baseline scores on the WAI-SR factors [21, 22].

The results reflect that TA depends on concordance (i.e., agreement), which determines the timing and duration of treatment. The quality of these interactions influences the extent to which TA develops between patient and practitioner. This underscores the potential adverse effects of placing patients on a waitlist, which may hinder the formation of TA. Nevertheless, the study demonstrated that TA can still be achieved despite a waitlist if sufficient time and frequent treatments are provided. Similar findings have been observed in studies employing fMRI hyper scanning, which highlighted specific brain regions such as the temporoparietal junction (TPJ) [30, 31], anterior insula (aINS) [31], dorsolateral and ventrolateral prefrontal cortex (dlPFC and vlPFC) [30, 32], and primary (S1) and secondary (S2) somatosensory areas [32]. These studies showed that TA influences vary based on whether patients had prior interactions with practitioners before hyperscanning. Participants with previous interactions characterized by social engagement (i.e., practitioner presence) and non-verbal cues (i.e., positive facial expressions, posture, gestures) [30]. Clinical intake (i.e., audio/video) [31, 32] exhibited stronger neural activations and self-reported experiences of empathy, rapport, and mutual trust, leading to lower pain levels.

Additionally, a study employing high temporal resolution EEG during patient-practitioner interactions found that behaviors categorized as “augmented” (i.e., warm, attentive relationships with active listening and personalized content) significantly impacted treatment outcomes. In contrast, “limited” interactions (i.e., neutral and impersonal with distracted clinicians) did not foster the same positive effects [34]. This suggests that TA, characterized by empathy, mutual trust, and warmth, enhances treatment adherence and clinical outcomes while alleviating chronic pain.

The present study’s hierarchical linear regression analyses demonstrated that treatment conditions significantly predicted pain levels and mediated relationships between TA and patient outcomes. Notably, WAI-SR factors, especially the Goal component, were associated with improved physical functioning in both groups over the six months. Data collected at the two-month mark indicated that the biweekly acupuncture group, which commenced treatment immediately, exhibited a higher Task component than the waitlist group, which was predictive of lower pain levels at six months. This finding aligns with the understanding that the waitlist group had not begun treatment; hence, no task formation could occur. The data emphasizes the importance of concordance (i.e., agreement) for achieving positive clinical outcomes. When the patient-practitioner relationship aligns, this concordance significantly influences their bond, reducing anxiety levels and patient stress relief.

### 4.1 Limitations

While the current analysis offers valuable insights, it may only partially capture the real-world effects of TA in populations beyond veterans diagnosed with GWI. However, existing evidence supports the influence of TA on clinical outcomes in populations with similar symptoms. Future research should explore the relationship between TA development, formation, and clinical outcomes in more diverse and representative samples from the general population. Additionally, this analysis lacked sufficient reflection of practitioner TA variables. Data from both sides of the patient-practitioner relationship are crucial for a comprehensive understanding of TA components. This information will enhance our understanding of whether TA formation, development, and outcomes are influenced by the practitioner’s experience or the dynamics of the TA itself.

### 4.2 Conclusions

This study illustrates that the development and formation of TA are essential for enhancing patient-practitioner relationships, facilitating patient engagement, and improving outcomes through tasks, goals, and bonds. Concordance is fostered when patients feel heard, understood, and supported, leading to positive measurable mediators such as empathy, rapport, and mutual trust. More evidence-based research is needed to explore TA with complementary medicine therapies, including acupuncture, massage, herbal medicine, and dietary therapy. Training for practitioners in achieving TA is critical, as it is vital for effective patient care and enhancing practitioner experiences. Ultimately, TA promotes higher adherence to treatment and alleviates symptoms (e.g., pain), which should be prioritized throughout all stages of the patient-practitioner relationship.

## 5. Declarations

The data was compiled from the 3.5-year US Army-funded RCT supported by the Office of the Assistant Secretary of Defense for Health Affairs through the Gulf War Illness Research Program under Award No. W81XWH-14-1-0533 and Grant number GW130028. The 3.5-year US Army-funded RCT was approved by the New England Institutional Review Board (IRB # 09-204). There were no awards, grants, funding, or IRB review boards for this TA review study. Opinions, interpretations, conclusions, and recommendations are those of the authors and are not necessarily endorsed by the Department of Defense.

## Data Availability

All data produced in the present work are contained in the manuscript.

## Acknowledgments

The original randomized clinical trial (RCT) that supplied the data for this study was supported by the Office of the Assistant Secretary of Defense for Health Affairs through the Gulf War Illness Research Program under Award No. W81XWH-14-1-0533 and approved by the New England Institutional Review Board (IRB # 09-204). Opinions, interpretations, conclusions, and recommendations are those of the author and are not necessarily endorsed by the Department of Defense.

